# Association Between Clinical Outcome Measures and Sonomyography-Derived Metrics in Individuals with Spinal Cord Injury

**DOI:** 10.64898/2026.07.27.26358334

**Authors:** Manikandan Shenbagam, Nikhil Chowdhary, Priyanka Vijay, Chitra Kataria, Biswarup Mukherjee

**Affiliations:** Centre for Biomedical Engineering, Indian Institute of Technology Delhi, New Delhi, 110016, Delhi, India.; Institute of Rehabilitation Services, Indian Spinal Injuries Centre Hospital, New Delhi, New Delhi, 110070, Delhi, India.; Department of Biomedical Engineering, All India Institute of Medical Sciences, New Delhi, 110029, Delhi, India.

**Author notes:** Contributing authors; priyanka.

**Keywords:** spinal cord injury, sonomyography, muscle activity sensing, upper extremity function

## Abstract

**Background:** To examine the association between ultrasound-based muscle activity detection, or sonomyography (SMG)-derived metrics, and clinical measures of upper-extremity function in individuals with cervical spinal cord injury (cSCI), and to evaluate SMG-based trajectories derived directly from muscle activity as a muscle-level assessment compared to conventional kinematic approaches.

**Methods:** Eight individuals with cSCI (n = 8; American Spinal Injury Association Impairment Scale grades A–C; injury levels C5–C6) participated. Participants performed a wrist tenodesis–based target achievement task while SMG data were collected. SMG-derived metrics were correlated with performance-based upper extremity function assessed using the Jebsen Taylor Hand Function Test (JTHFT) and self-reported function assessed using the Capabilities of Upper Extremity Questionnaire (CUE-Q). Associations were quantified using distance correlation (dCorr).

**Results:** Strong associations between SMG-derived metrics and clinical measures were observed. Movement Arrest Period Ratio (MAPR) showed the strongest association with JTHFT performance (*d*_Corr_ = 0.75), while Time to Peak Velocity (TTPV) demonstrated a moderate association (*d*_Corr_ = 0.62). Rate of Change of Acceleration (ROCAcc) showed a strong correlation with CUE-Q scores (*d*_Corr_ *≈* 0.70), and spectral arc length (SAL) showed moderate correlations (*d*_Corr_ *≈* 0.66).

**Conclusions:** SMG-derived metrics show meaningful associations with both performance-based and self-reported measures of upper-extremity function in individuals with cSCI. These findings suggest that SMG metrics can serve as objective tools to complement clinical assessments for tracking functional status and recovery. Larger studies are needed to confirm these observations.

## 1 Background

Cervical spinal cord injury (cSCI) affects the normal functioning of motor, sensory, and autonomic pathways below the site of injury, leading to significant difficulties in voluntary movement and sensation. Around 60% of individuals with spinal cord injury (SCI) experience injuries at the cervical level, which often results in tetraplegia and reduced function of the upper limbs, trunk, and lower limbs [1]. Globally, the occurrence of SCI varies widely in terms of causes and clinical presentation, with falls and road traffic accidents being the most common reasons [2]. Reported incidence rates range from 13–163.4 per million in developed countries and 13–220 per million in non-developed countries, while prevalence estimates range from 490–526 per million in developed countries and approximately 440 per million in non-developed countries, respectively [3]. Traumatic SCI is more frequently motor-complete, classified as American Spinal Injury Association Impairment Scale (AIS) grades A–B, whereas non-traumatic SCI more commonly presents as motor-incomplete AIS grades C–D [4]. Among individuals with tetraplegia, recovery of upper extremity function is consistently ranked as the highest rehabilitation priority [5, 6]. Functional use of the arms and hands is essential for performing activities of daily living, maintaining independence, and improving overall quality of life [7, 8]. Following cSCI, impairment of upper limb function commonly arises from partial or complete paralysis of intrinsic and extrinsic hand and forearm muscles, leading to significant difficulties with grasping, object manipulation, and coordinated hand movements. At the same time, even small neurological improvements, such as recovery at a single cervical segment, can lead to noticeable functional benefits. According to Consortium for Spinal Cord Medicine [9], this can significantly impact independence in fundamental functions such as feeding, grooming, and mobility. Therefore, improving upper limb function is a primary goal in rehabilitation programs, clinical research, and various organisations working in the area of SCI.

Clinical outcome assessments (COAs) are often used to measure upper limb function in individuals with cSCI, including clinician-rated scales, performance-based tests, and self-reported questionnaires [10]. While these methods are essential for clinical decision-making, they are often performed at specific time periods and mainly focus on task completion, the strength involved, or the patient’s perceived ability. Because of this, they don’t completely capture the underlying strategies for motor control and movement quality and how the performance of the upper limb varies in everyday activities. In addition, many commonly used assessments are not sensitive enough to detect small or compensatory changes in movement, especially during the early stages of recovery or in individuals who still have some voluntary control but with impairments. As a result, improvements in real-world upper limb performance may not be adequately captured by conventional assessment scores alone [11].

To address these limitations, physiological sensing modalities have been explored to provide objective quantification of muscle activity and movement characteristics. Surface electromyography (sEMG) has been widely used for intent detection and gesture recognition [12, 13]; however, in individuals with SCI, sEMG signals are often weak and inconsistent due to reduced voluntary activation, low signal-to-noise ratios, crosstalk, motion artefacts, and sensitivity to electrode placement and skin conditions [14]. Although high-density sEMG has demonstrated improved detection of spared motor unit activity, even in motor-complete SCI [15, 16], its reliance on specialised electrode arrays and complex instrumentation limits its practicality for long-term or wearable applications. Ultrasound-based muscle activity sensing or sonomyography (SMG) has emerged as a promising alternative by capturing mechanical muscle deformations associated with contraction, rather than electrical activity. SMG enables access to both superficial and deep muscle compartments with higher spatial resolution and improved signal robustness compared to sEMG [17, 18]. These characteristics make SMG particularly well suited for individuals with SCI, including those with discomplete SCI, where voluntary muscle activity may be subtle or difficult to detect using conventional electrical methods [19–21]. Previous studies have demonstrated that ultrasound based systems can achieve strong performance in tasks such as gesture classification, force estimation, and proportional control of wrist and hand movements, while enabling real-time and wearable implementations [22–24]. In our previous work [19], we developed a human–machine interface (HMI) that captured muscle activity using ultrasound imaging and mapped the derived SMG signals to one-dimensional cursor movement. Participants were required to achieve visually presented targets using ultrasound-derived muscle activity, enabling continuous assessment of motor performance. From the resulting cursor trajectories, quantitative outcome measures were extracted to characterise movement quality. Similar target acquisition tasks (TACs) have been widely employed using inertial measurement units (IMUs) to control cursor motion, with kinematic trajectories used to derive measures of movement smoothness, speed, and control [25–27]. These trajectory-based metrics have been shown to sensitively reflect motor control integrity and functional impairment. In particular, sensor-derived movement quality metrics such as smoothness have emerged as robust indicators of neuromotor function and recovery. Prior studies have reported significant associations between kinematic features, including smoothness, peak velocity, and movement time, and established COAs in individuals with neurological impairments. Such approaches provide an opportunity to complement conventional assessments by offering objective, continuous, and high-resolution characterisation of upper extremity function. Wearable sensing technologies, in particular, hold promise for bridging the gap between laboratory-based assessments and real-world functional performance in clinical and community settings.

The objective of this study was to investigate the relationship between SMG-derived metrics and upper extremity function as assessed by established COAs. This study was conducted in a diverse cohort of individuals with acute, subacute, and chronic cSCI undergoing inpatient rehabilitation. By examining the associations between sonomyography-based metrics and both performance-based and self-reported COAs, this work aims to evaluate the clinical relevance of SMG as an objective tool for quantifying upper extremity motor function after cSCI.

## 2 Materials and Methods

### 2.1 Participants

The participants and data used in this study have been previously described in our earlier work [19]. Briefly, eight individuals with cSCI (mean age: 29 *±* 12 years) were recruited. Written informed consent was obtained from all participants, and the study was approved by the Institute Ethics Committees (IEC) of Indian Spinal Injuries Centre (ISIC), New Delhi, and Indian Institute of Technology, Delhi (ISIC/RP/2022/05 and P021/P050, respectively). Participants were selected based on predefined eligibility criteria following a detailed neurological examination (Table 1). Demographic information and clinical assessment scores for all participants have been detailed in Supplementary Table 1.

**Table 1:** Eligibility Criteria.

| Category | Criteria |
| --- | --- |
| Inclusion Criteria | Cervical spinal cord injury (C5–C6), ASIA Impairment Scale grades A, B, or C and D<br>Age between 18 and 60 years |
| Exclusion Criteria | Acute medical instability or severe visual impairment<br>Upper-limb deformity affecting task performance<br>History of psychiatric, neurological, or musculoskeletal disorders unrelated to SCI<br>Refusal or inability to provide informed consent or follow experimental commands<br>Presence of diabetes or chronic pain conditions |

### 2.2 Procedures

Participants were seated comfortably during the experiment, with the dominant arm supported on a platform such that the elbow rested directly beneath the shoulder on a cushioned armrest as shown in Figure 1(a). This standardized positioning was used to minimize fatigue and to ensure consistency across participants during data collection. Muscle activity related to the tenodesis grasp was measured using clinical ultrasound imaging system (Model: PT60 A, Samsung Electronics, Suwon, South Korea). A high-frequency linear ultrasound probe (Model: LV 7.5/60/128 Z-2, Samsung Electronics, Suwon, South Korea) was placed over the dorsal aspect to assess major extensor muscle groups, secured using a custom made holder with Velcro straps. Ultrasound gel (Sono-care transmission gel, Hi-tech Surgical & Fitness, Ahmedabad, India) was applied to ensure adequate contact between the probe and the skin. The probe was positioned transversely approximately 5 cm distal to the olecranon process to image the primary forearm extensor muscles involved in tenodesis grasp, including the extensor carpi radialis longus, extensor carpi radialis brevis, and extensor carpi ulnaris. Ultrasound images were streamed in real time to a custom interface at an average frame rate of approximately 20 frames per second. The collected ultrasound images were stremed to streamed in real time to a custom-developed MATLAB interface (Version 2022a, MathWorks Inc., Natick, MA, USA) running on a laptop computer (Intel Core i7–7700HQ, 32 GB RAM, 4 GB NVIDIA GeForce GTX 1040Ti) using a USB-based video capture device (Model: Corsair Elgato Game Capture HD60S+, Kowloon, Hong Kong).

**Fig. 1:**
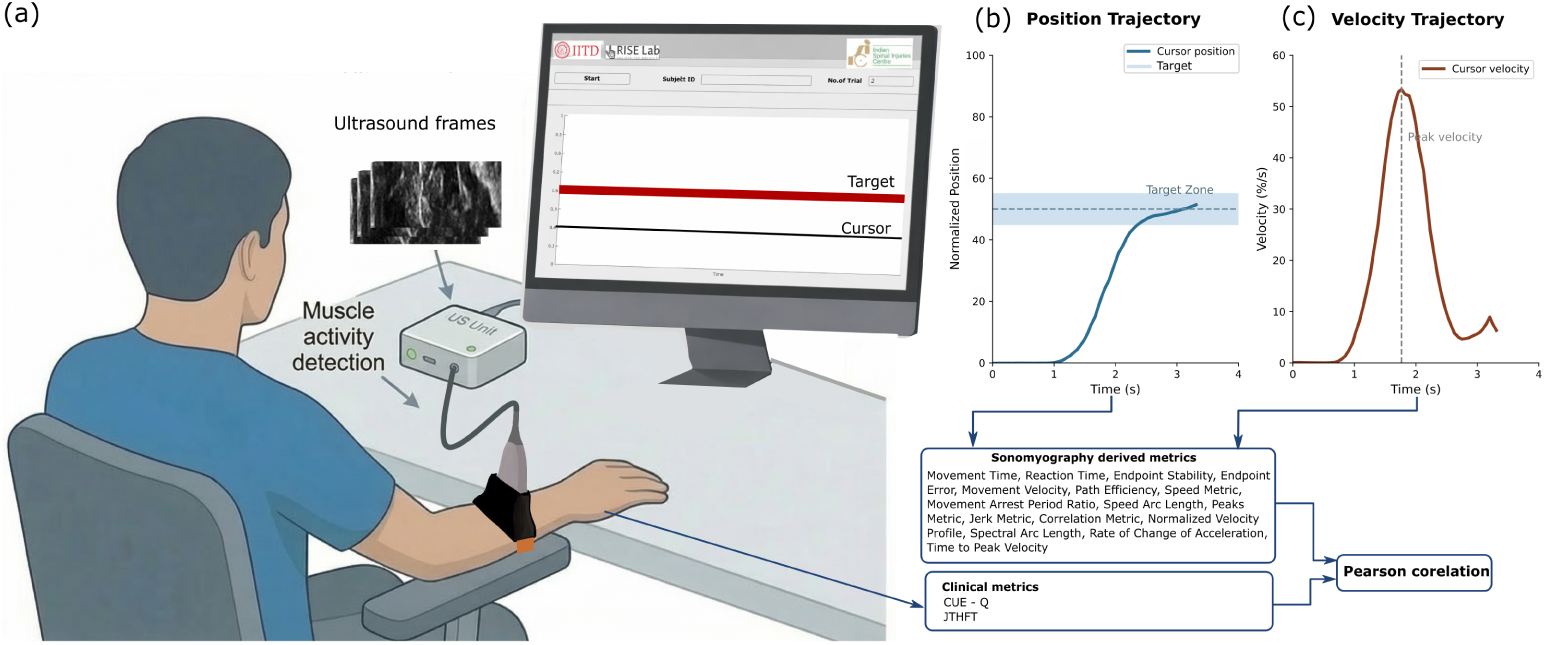
(a) Experimental setup showing individuals with SCI instrumented with an ultrasound probe on the forearm. A clinical ultrasound machine captures muscle activity as brightness-mode (B-mode) images. A MATLAB-based algorithm computes the correlation between incoming and reference B-mode images and maps this signal to cursor movement, which is displayed on the screen. (b) Representative position trajectory showing the normalized cursor position approaching and stabilizing within the target region over time; the shaded region indicates the target. (c) Velocity trajectory illustrating the temporal evolution of movement dynamics during target acquisition.

### 2.3 Calibration of Wrist Movement Range

Before the experimental task, each participant completed a calibration phase to determine their individual wrist movement range. Participants were instructed to repeatedly perform the tenodesis movement with maximum comfortable wrist extension followed by complete relaxation for approximately 30 seconds. Ultrasound images were recorded and stored in both the fully extended and fully relaxed positions to serve as reference states. This calibration enabled muscle activity signals to be normalized to each participant’s own functional range, thereby ensuring that task difficulty was tailored properly as per the individual.

### 2.4 Target-Achievement Task

After the calibration step, participants carried out a target-reaching task using a cursor displayed on the screen. The vertical movement of the cursor was controlled using the muscle activity signal obtained from ultrasound, as shown in Figure. 1(a). Wrist extension moved the cursor upward and wrist relaxation moved it downward. They were asked to move the cursor from a resting position to a given target using a tenodesis grasp. A total of nine target levels were used, ranging from 10% to 90% of each participant’s calibrated movement range, and these were presented in a random order. Each target had an allowable margin of ±5%. Participants were given up to 10 seconds to reach the target, and a trial was marked successful if they could hold the cursor within the target region continuously for at least 1.5 seconds. After every trial, the cursor automatically returned to the resting position so that each attempt started under the same conditions. To reduce fatigue and the risk of muscle strain, the targets were limited to 90% of the calibrated range. An auditory cue was used to start each trial. Participants were instructed not to look at their hand during the task, encouraging more reliance on their sense of movement rather than visual feedback.

### 2.5 Ultrasound Signal Processing and Cursor Control

A sonomyography signal was obtained by processing ultrasound images acquired in real time during the task, reflecting changes in muscle morphology. [28]. This signal was normalised using participant-specific calibration values. To address gradual variations, including minor changes in probe placement or posture, the normalisation limits were continuously updated during the task. This approach ensured stable and consistent cursor control throughout the course of the experiment. The data analysed in this study were collected as part of our previously published work using the same experimental protocol and participant cohort.

### 2.6 Clinical Assessment and Data Collection

Participants completed the Jebsen Taylor Hand Function Test (JTHFT) [29] and the Capabilities of Upper Extremity Questionnaire (CUE-Q) [30] to assess performance-based and self-reported upper-extremity function, respectively. The JTHFT is a standardised assessment of functional hand use with established reliability, validity, and responsiveness in neurological populations. A composite JTHFT time score was computed by summing the completion times across all subtests performed with the tested upper extremity, with lower scores indicating better hand function. Self-reported upper extremity function was evaluated using the CUE-Q, in which participants rated the difficulty of performing daily tasks on a Likert scale ranging from 0 (unable to perform) to 7 (no difficulty). A total CUE-Q score was obtained by summing the scores across all questionnaire items. Detailed descriptions of the Jebsen–Taylor Hand Function Test and Capabilities of Upper Extremity Questionnaire are provided in Supplementary Table 2 and 3.

### 2.7 SMG-Derived Performance Metrics

SMG-derived performance and movement quality metrics were computed from successful trials and used for correlation analysis with clinical outcome measures (Figure 2). Reaction time, movement time, endpoint stability, endpoint error, maximum velocity, and path efficiency were adopted from trajectory-based analyses previously developed for SMG-based cursor control in individuals with SCI [19]. The remaining kinematic and smoothness metrics, including movement smoothness quantified using the Spectral Arc Length (SPARC) metric, were derived from established movement analysis frameworks in motor control and neurorehabilitation literature and were originally validated in stroke populations [25]. During the target achievement task, the cursor position was recorded as a function of time, producing a continuous position trajectory **p**(*t*), as illustrated in Figure. 1(b). The corresponding velocity trajectory **v**(*t*) was obtained by differentiating the position signal, and its magnitude was used to compute scalar speed as shown in Figure. 1(c):

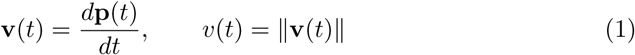

**Fig. 2:**
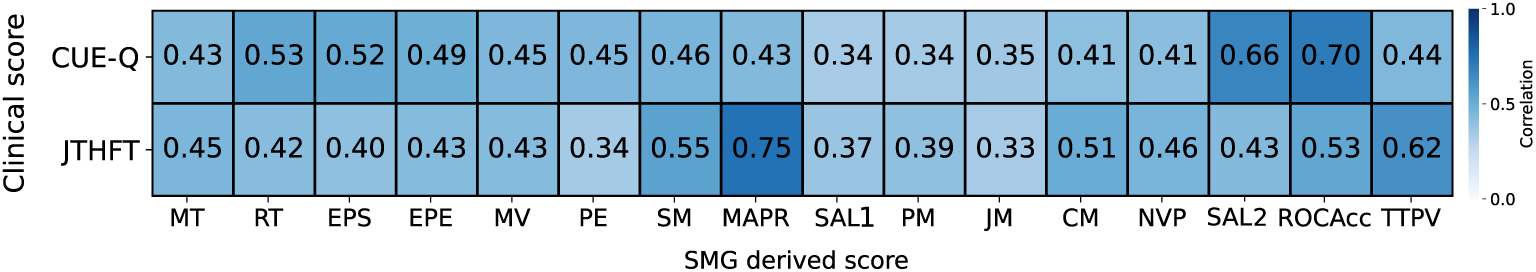
Correlation heatmap illustrating the pairwise relationships between clinical outcome measures and sonomyography (SMG)-derived movement quality metrics. Rows correspond to clinical scores, including Level of Injury (LI), Capabilities of Upper Extremity Questionnaire (CUE-Q), and Jebsen–Taylor Hand Function Test (JTHFT), while columns represent SMG-derived metrics: Movement Time (MT), Reaction Time (RT), Endpoint Stability (EPS), Endpoint Error (EPE), Movement Velocity (MV), Path Efficiency (PE), Speed Metric (SM), Movement Arrest Period Ratio (MAPR), Speed Arc Length (SAL1), Peaks Metric (PM), Jerk Metric (JM), Correlation Metric (CM), Normalized Velocity Profile (NVP), Spectral Arc Length (SAL2), Rate of Change of Acceleration (ROCAcc), and Time to Peak Velocity (TTPV). Each cell reports the distance correlation coefficient between the corresponding clinical score (row) and SMG-derived metric (column), with color intensity indicating the strength of the association. Moderate to strong correlations were seen across different metrics. Among them, MAPR showed the most consistent and strongest relationship with the clinical measures. Metrics related to smoothness and coordination also showed moderate levels of association with the clinical outcomes.

Here, **p**(*t*) denotes the cursor position vector at time *t*, **v**(*t*) denotes the velocity vector, and *v*(*t*) represents the scalar speed. The signals were sampled at frequency *f_s_*over the total trial duration *T*. The following metrics were extracted from successful trials only, defined as those in which the cursor entered and remained within the target bounds for the prescribed dwell duration.

#### 2.7.1 Reaction Time (RT)

Reaction time represents the latency between the presentation of the target and the initiation of cursor movement. In the position trajectory, this corresponds to the interval between target onset and the first detectable deviation of the cursor from the resting baseline (typically defined as a 5 % change in position threshold) [31].

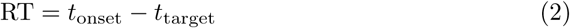

where *t*_target_ is the time of target presentation and *t*_onset_ is the time at which movement initiation is detected.

#### 2.7.2 Movement Time (MT)

Movement time represents the duration required for the cursor to travel from movement onset to the instant it first reaches the target region.

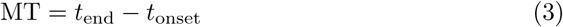

where *t*_end_ denotes the time at which the cursor first enters the target bounds.

#### 2.7.3 Endpoint Stability (EPS)

Endpoint stability quantifies the variability of the cursor position during the dwell phase after target acquisition [32].

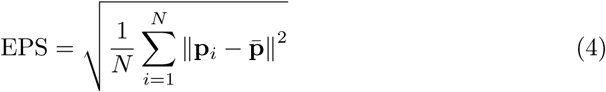

where **p***_i_*are cursor samples recorded during the dwell interval, **p̄** is the mean cursor position during this period, and *N* is the number of samples in the dwell phase.

#### 2.7.4 Endpoint Error (EPE)

Endpoint error measures the spatial accuracy of the cursor by computing the distance between the final cursor position and the target location. [32].

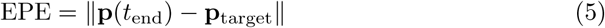

where **p**_target_ represents the target position.

#### 2.7.5 Maximum Velocity (MV)

Maximum velocity corresponds to the peak value of the speed profile obtained from the velocity trajectory.

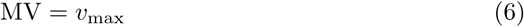

where *v*_max_ = max*_t∈_*_[0*,T* ]_ *v*(*t*).

#### 2.7.6 Path Efficiency (PE)

Path efficiency evaluates the straightness of the movement trajectory by comparing the actual path length to the ideal straight-line distance between the starting point and the target [33].

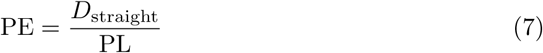

where *D*_straight_ = ||**p**_target_ *−* **p**(*t*_onset_)|| is the straight line distance and PL is the actual path length defined as

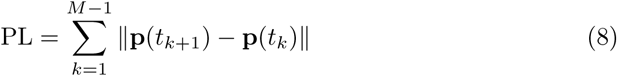

with *M* denoting the total number of samples between movement onset and target entry.

#### 2.7.7 Speed Metric (SM)

The speed metric characterizes the uniformity of the velocity profile by comparing the mean speed to the peak speed [34].

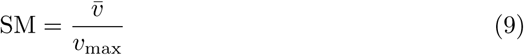

where 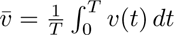 is the mean speed over the movement duration.

#### 2.7.8 Movement Arrest Period Ratio (MAPR)

MAPR quantifies the proportion of time during which the cursor is actively moving [35].

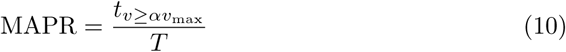

where *α* is a predefined threshold and *t_v≥αv_*_max_ represents the cumulative time for which *v*(*t*) exceeds *αv*_max_.

#### 2.7.9 Speed Arc Length (SAL_1_)

Speed arc length is a time-domain smoothness metric computed from the normalized velocity trajectory [27].

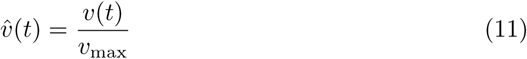

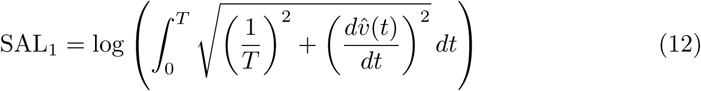

#### 2.7.10 Peaks Metric (PM)

The peaks metric represents the number of prominent local maxima in the velocity profile and reflects the presence of corrective submovements [36].

#### 2.7.11 Jerk Metric (JM)

The jerk metric quantifies movement smoothness using the squared third derivative of position.

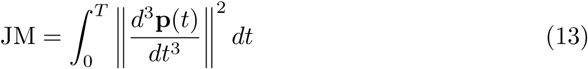

#### 2.7.12 Correlation Metric (CM)

The correlation metric evaluates the similarity between the measured velocity profile and an ideal minimum-jerk velocity trajectory [37].

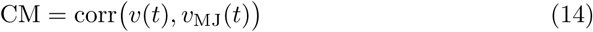

where *v*_MJ_(*t*) denotes the velocity predicted by the minimum-jerk model [19].

#### 2.7.13 Normalized Velocity Profile (NVP)

The normalized velocity profile is obtained by scaling the velocity trajectory by its peak value:

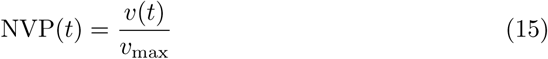

#### 2.7.14 Spectral Arc Length (SAL_2_)

Spectral arc length is a frequency-domain smoothness metric computed from the magnitude spectrum of the velocity signal [27].

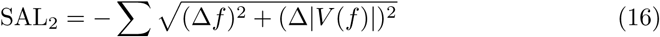

where *V* (*f*) denotes the Fourier transform magnitude of *v*(*t*).

#### 2.7.15 Rate of Change of Acceleration (ROCAcc)

The rate of change of acceleration captures rapid fluctuations in acceleration [32].

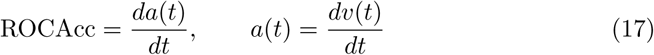

#### 2.7.16 Time to Peak Velocity (TTPV)

Time to peak velocity describes the temporal organization of the movement by indicating when the maximum speed occurs relative to the total movement duration.

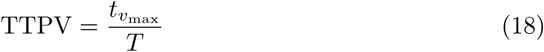

where *t_v_*_max_ denotes the time at which the peak velocity *v*_max_ occurs.

### 2.8 Statistical Analysis

Statistical analysis was performed to examine the association between SMG-derived metrics obtained during the target-achievement task and clinical measures of upper extremity function. Specifically, outcome metrics extracted from the sonomyography trajectories were compared with performance-based scores from the JTHFT and self-reported scores from the CUE-Q. Distance correlation (dCorr) analysis was used to quantify these associations across the participant cohort. This method was selected because it is sensitive to both linear and nonlinear relationships and does not require assumptions about the underlying form of the association. Distance correlation values are nonnegative, with a value of zero indicating no association and higher values indicating stronger relationships. A threshold of 0.50 was used to identify SMG-derived metrics that showed a strong association with clinical measures.

## 3 Results

Distance correlation analysis was performed to examine associations between sonomyography-derived metrics and clinical measures of upper extremity function.

### 3.1 Association Between SMG-Derived Metrics and Clinical Outcome Measures

Fig. 2 summarizes the strength of association between SMG-derived metrics and established clinical outcome measures, including self-reported upper extremity function (CUE-Q) and performance-based upper extremity function (JTHFT). Across metrics, moderate to strong correlations were observed, indicating that ultrasound-derived measures of movement quality are meaningfully related to clinical function. Among the SMG-derived metrics, Movement Arrest Period Ratio (MAPR) demonstrated the strongest association with JTHFT performance (*d*_Corr_ = 0.75). Time to Peak Velocity (TTPV) also showed a moderate to strong association with JTHFT (*d*_Corr_ = 0.62), along with other coordination-related metrics such as speed metric (SM; *d*_Corr_ = 0.55) and correlation metric (CM; *d*_Corr_ = 0.51). Associations with self-reported function were also evident. Rate of Change of Acceleration (ROCAcc) exhibited the strongest correlation with CUE-Q scores (*d*_Corr_ = 0.70), followed by spectral arc length at higher target levels (SAL2; *d*_Corr_ = 0.66). Several temporal and endpoint-related metrics, including reaction time (RT) and endpoint position stability (EPS), showed moderate correlations with CUE-Q (up to *d*_Corr_ *≈* 0.53). In contrast, metrics reflecting simpler temporal features generally demonstrated weaker associations. These findings show that SMG-based measures capturing elements like movement smoothness, coordination, and overall movement pattern are more strongly associated with meaningful changes in upper limb function than basic task-level measures.

### 3.2 Target-Level Dependence of Correlation With Self-Reported Function (CUE-Q)

Fig. 3 (a) illustrates the relationship between SMG-derived metrics and self-reported upper-limb function, measured using the CUE-Q, across different target levels. The dcorr values varied across different targets and did not consistently increase with task difficulty. Certain SMG metrics showed higher correlations at lower levels of the target. For instance, ROCAcc had a peak at Target 10, and SAL2 showed higher correlations at some low to mid targets. This indicates that there is a close link between how well a perceived functional ability is and how well they perform controlled, small amplitude movements of tenodesis.

**Fig. 3:**
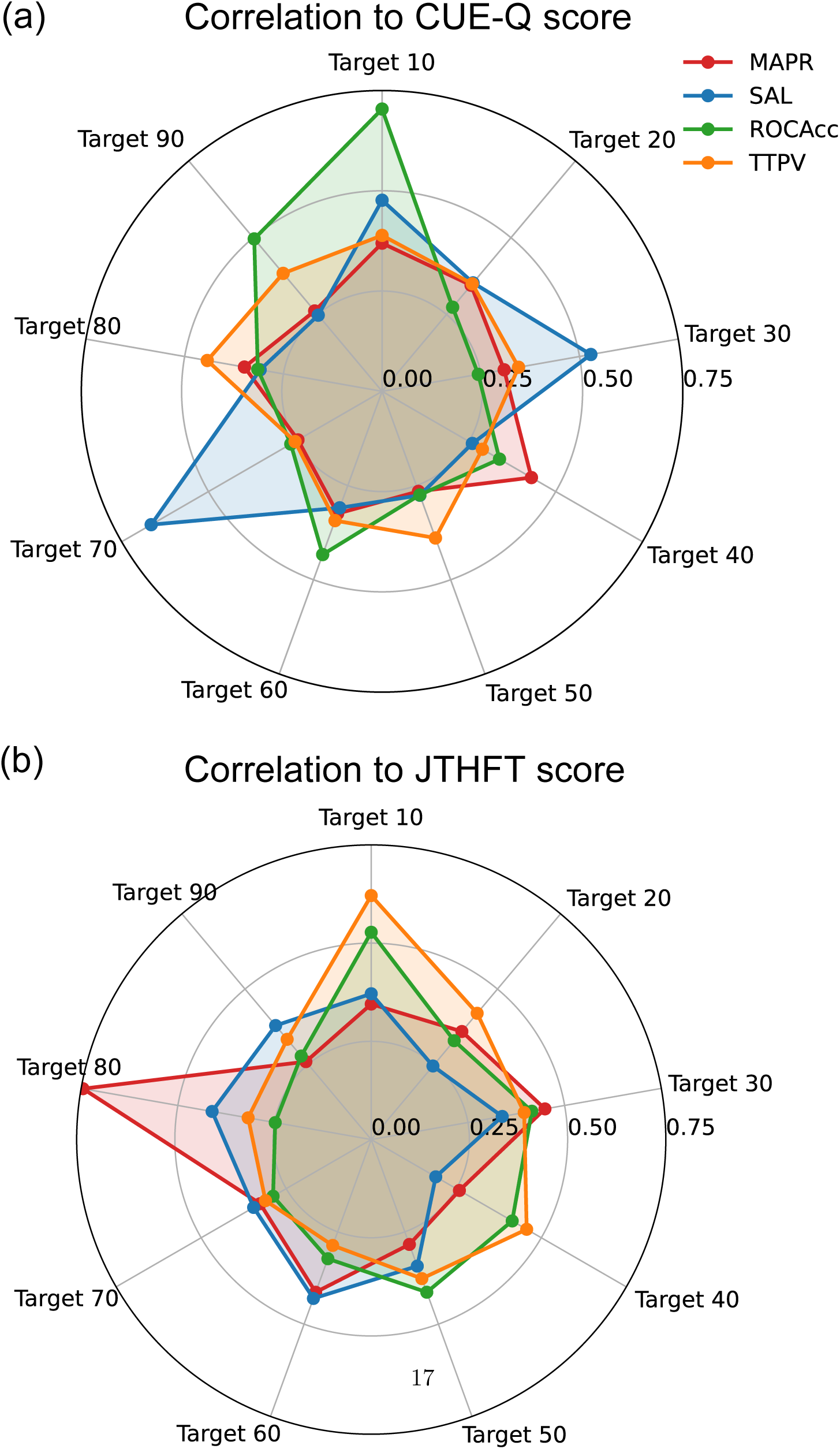
Target-wise correlation between clinical scores and SMG-derived metrics. (a) Radar plots illustrating the target-wise correlation between the Capabilities of Upper Extremity Questionnaire (CUE-Q) and sonomyography (SMG)–derived metrics across different targets achieved during the experiment. The SMG-derived metrics include MAPR (Movement Arrest Period Ratio), SAL2 (Spectral Arc Length), ROCAcc (Rate of Change of Acceleration), and TTPV (Time to Peak Velocity). (b) Radar plots illustrating the target-wise correlation between the Jebsen–Taylor Hand Function Test (JTHFT) scores and the same set of SMG-derived metrics across targets.

The correlations were less consistent at higher target levels and varied across different metrics. SAL2 demonstrated a noticeable peak at Target 70, while MAPR and TTPV showed moderate correlations at higher targets without a consistent pattern across all levels. The lower correlations at some intermediate targets suggest that self reported function may be more influenced by how the movement is done than by how hard the task is overall. These findings suggest that CUE-Q scores capture both fine motor control at lower targets and certain aspects of movement execution at higher targets. This emphasizes the importance of analysing SMG-based metrics across different task conditions instead of focusing solely on high effort movements.

### 3.3 Target-Level Dependence of Correlation With Performance-Based Hand Function (JTHFT)

Fig. 3(b) illustrates how SMG-based metrics are related to performance-based hand function measured using the JTHFT across different target levels. The dcorr values indicate that SMG metrics have meaningful associations with JTHFT scores at both lower and higher targets. For example, TTPV demonstrated a strong correlation at the lowest target level (Target 10), indicating that movement time during small tenodesis movements is closely linked to hand function. MAPR and SAL2 also demonstrated moderate correlations at lower targets, indicating their ability to capture movement interruptions and trajectory features during controlled actions.

At higher target levels, MAPR showed a clear peak in correlation at Target 80, which suggests that movement stopping characteristics become more important as task difficulty increases. TTPV and SAL2 continued to show moderate correlations at some higher targets, while ROCAcc showed relatively consistent but smaller associations across all levels. Even though the correlations did not steadily increase with target level, the presence of strong relationships at both low and high targets suggests that SMG-based metrics are able to capture clinically meaningful aspects of hand function over a wide range of task conditions. Overall, these findings highlight that SMG-based metrics can reflect both subtle motor control issues during easier tasks and performance related difficulties during more challenging movements in individuals with cSCI.

Taken together, these findings demonstrate that ultrasound-derived measures of movement quality during a tenodesis-based task are significantly associated with both self-reported and performance-based clinical assessments, especially under higher task demands. This highlights their potential clinical relevance for quantifying upper extremity motor function after cSCI.

## 4 Discussion

This study investigated the relationship between ultrasound-based SMG–derived metrics and established clinical measures of upper extremity function in individuals with cSCI. Several SMG-derived features were found to have moderate to strong associations with both performance-based (JTHFT) and self-reported (CUE-Q) outcomes across various target conditions. This suggests that muscle-level movement characteristics captured by SMG are meaningfully related to functional upper extremity ability. These associations were target-dependent but not monotonic, with clinically relevant correlations observed at both lower and higher target levels. These findings suggest that SMG-based metrics may serve as useful objective indicators for assessing upper limb function in individuals with cSCI.

### 4.1 Movement Intermittency as a Marker of Functional Impairment

Among the evaluated metrics, MAPR showed strong and target specific correlations with COAs of upper extremity function. Higher MAPR values, which indicate more frequent movement pauses and fragmented execution [25],were associated with poor performance on the JTHFT and lower self-reported upper extremity capability. These associations were particularly evident at higher target levels, notably around Target 80. This indicates that the significance of movement intermittency becomes more relevant when tasks demand greater amplitude and prolonged control. These findings align with previous observations that individuals with neurological injury frequently employ segmental and corrective movement strategies, likely due to impaired feedforward control and reduced motor unit recruitment. Overall, the observed sensitivity of MAPR across targets highlights it as a salient indicator of impaired motor control after cSCI.

### 4.2 Characteristics of Movement Execution and Smoothness

Metrics associated with movement timing and acceleration, such as TTPV and ROCAcc, showed meaningful associations with both clinical outcome measures across various target levels. TTPV, in particular, demonstrated strong correlations at lower targets. This suggests that the timing of small tenodesis movement is closely linked to actual hand function. ROCAcc demonstrated moderate and fairly consistent correlations across targets, indicating that the regulation of acceleration influences functional ability under different task demands. Overall, these findings suggest that SMG-based temporal features are sensitive to impairments in motor planning, force production, and coordination, even when the movements are not very complicated. Metrics related to smoothness, such as SAL2, exhibited varying relationships with clinical scores depending on the target level, with notable peaks at certain higher targets. Reduced movement smoothness is often indicative of impaired motor control and an increased reliance on corrective adjustments. The findings indicate that SMG-based smoothness metrics can capture underlying neuromuscular control issues and offer additional insights beyond traditional COAs, which are primarily focused on task completion or perceived ability.

### 4.3 Influence of Task Demand on Clinical Associations

The relationship between metrics derived from SMG and clinical outcome measures demonstrated variability across different target levels and did not simply increase with task difficulty. There were meaningful correlations at both the lower and higher targets. At the lower target levels, involving small and controlled movements, they were more sensitive to issues related to timing and coordination. On the other hand, higher target levels highlighted problems such as movement interruptions and difficulties in execution. This kind of irregular trend emphasizes the importance of evaluating performance across a range of task conditions, instead of focusing solely on challenging movements.

Differences were also observed between the two clinical measures used. The performance-based JTHFT had stronger associations with SMG metrics at certain low and high target levels, thereby demonstrating its sensitivity to deficits in the actual execution of movement. Conversely, the CUE-Q, on the other hand, a self-reported measure, exhibited greater variability across targets, reflecting its subjective nature and its focus on perceived ability. Together, these findings suggest that both clinical assessments provide valuable yet distinct information, and that metrics based on SMG are able to capture movement characteristics related to both actual performance and perceived functional ability.

### 4.4 Clinical Implications and Future Directions

The results of this study indicate that SMG-based metrics may serve as effective noninvasive and quantitative measures of upper extremity function in individuals with cSCI. These metrics, by capturing aspects such as movement interruptions, timing, and smoothness under different task conditions, provide additional insights beyond those obtained by standard COAs and may help in more sensitive detection of motor impairments.

Meaningful correlations were observed even at lower target levels, which indicates that subtle motor deficits can be identified without requiring participants to perform highly challenging tasks. This can also be very beneficial for individuals with severe impairments. Future work should examine the temporal variation of these metrics changing over time and assess their applicability across a broader range of functional tasks and varying levels of injury severity.

### 4.5 Limitations

The study had a relatively small number of participants, and as it was cross-sectional, it is not possible to tell if there are causal relationships or how these measures change over time. Moreover, the analysis was confined to specific target levels within a controlled experimental setup, which may not fully represent how movements occur in real life daily activities. Despite these limitations, the target specific correlations observed in this study offer useful evidence highlighting the clinical significance of SMG-based metrics.

## 5 Conclusions

This study demonstrates that SMG-derived metrics are meaningfully associated with measures of upper extremity function in individuals with cSCI. Metrics capturing movement intermittency, timing, acceleration, and smoothness showed moderate to strong correlations with both performance-based function, assessed using the JTHFT, and self-reported function, measured using the CUE-Q. Performance-based outcomes were particularly sensitive to execution related features such as movement arrest and timing, while self-reported function exhibited target specific associations with smoothness and acceleration related metrics. Importantly, these relationships were dependent on task condition but did not increase monotonically with target demand. Clinically meaningful associations were observed at both lower and higher target levels, indicating that SMG-derived metrics can capture subtle impairments during low amplitude movements as well as execution-related deficits during more demanding tasks. Collectively, these findings support the feasibility of SMG-derived metrics as objective, non-invasive markers of upper extremity motor control after SCI and provide preliminary evidence for their further development as digital biomarkers for functional assessment and rehabilitation monitoring.

## Declarations

### Ethics approval and consent to participate

All study participants provided informed consent, and the study protocol was approved by the Institutional Ethics Committee of the Indian Spinal Injuries Centre (ISIC), New Delhi, under approval number ISIC/RP/2022/05.

### Consent for publication

Written informed consent for publication of data and images (including de-identified TRA demographic information) was provided from all the participants.

### Availability of data and materials

The datasets generated and analysed during the current study are not publicly available due to absence of a formal data-sharing agreement with the participants at the time of data collection. De-identified data may be made available on reasonable request from the corresponding author, subject to institutional ethical approval.

### Competing requests

The authors declare that they have no competing interest.

### Authors’ contributions

MS designed the study, developed the data processing pipeline, led data collection for individuals with SCI, conducted all analyses, and was the primary author of the manuscript. NC contributed to the experimental setup and data collection. PV and CK provided clinical supervision, facilitated the recruitment of individuals with SCI, and guided the interpretation of the clinical findings. BM conceptualised the study, provided overall supervision, guided data interpretation, and critically revised the manuscript. All authors read and approved the final manuscript.

## Data Availability

All data produced in the present study are available upon reasonable request to the authors

